# Norwich COVID-19 Testing Initiative pilot: evaluating the feasibility of asymptomatic testing on a university campus

**DOI:** 10.1101/2020.09.22.20199455

**Authors:** T Berger Gillam, J Cole, K Gharbi, E Angiolini, T O Barker, P Bickerton, T Brabbs, J S Chin, E Coen, Sarah Cossey, R P Davey, R Davidson, A Durrant, D Edwards, N Hall, S Henderson, M Hitchcock, N Irish, J Lipscombe, G Jones, G Parr, S Rushworth, N Shearer, R Smith, N Steel

**Affiliations:** Health Services and Primary Care Research Group, University of East Anglia, Norwich, NR4 7TJ, United Kingdom; Genomics Pipelines, Earlham Institute, Norwich, NR4 7UZ; Scientific Training and Education, Earlham Institute, Norwich, NR4 7UZ; Communications, Earlham Institute, Norwich, NR4 7UZ; School of Computing Sciences, University of East Anglia, Norwich, NR4 7TJ. United Kingdom; John Innes Centre, Norwich, NR4 7UH; Earlham Institute, Norwich, NR4 7UZ; Faculty of Medicine and Health Sciences, University of East Anglia, Norwich, NR4 7TJ. United Kingdom; UEA Biosciences, University of East Anglia, Norwich, NR4 7TJ. United Kingdom; UEA Health and Social Care Partners, University of East Anglia, Norwich Research Park, Norwich NR4 7TJ, United Kingdom; University of East Anglia, Norwich, NR4 7TJ. United Kingdom; Faculty of Science, University of East Anglia, Norwich, NR4 7TJ. United Kingdom

## Abstract

**Background:** There is a high prevalence of COVID-19 in university-age students, who are returning to campuses. There is little evidence regarding the feasibility of universal, asymptomatic testing to help control outbreaks in this population. This study aimed to pilot mass COVID-19 testing on a university research park, to assess the feasibility and acceptability of scaling up testing to all staff and students.

**Methods:** This was a cross-sectional feasibility study on a university research park in the East of England. All staff and students (5,625) were eligible to participate. All participants were offered 4 PCR swabs, which they self-administered over two weeks. Outcome measures included: uptake; drop-out rate; positivity rates; participant acceptability measures; laboratory processing measures; data collection and management measures.

**Results:** 798 (76%) of 1053 who registered provided at least one swab. 687 (86%) provided all four. 792 (99%) of 798 who submitted at least one swab had all negative results. 6 participants had one inconclusive result. There were no positive results. 458 (57%) of 798 participants responded to a post-testing survey, demonstrating a mean acceptability score of 4.51/5, with 5 being the most positive.

**Conclusions:** Repeated self-testing for COVID-19 using PCR is feasible and acceptable to a university population.

## Introduction

Universities are considering methods of dealing with the transmission of COVID-19 when students return to campus. Student populations are likely to have a higher than average prevalence of infection (1) and in particular, a high rate of asymptomatic infection (2). This population is also highly mobile and more likely to have a large number of social contacts (3). It remains unclear how an outbreak might evolve on a university campus, but modelling studies suggest that students are highly interconnected, indicating significant potential for infectious disease transmission (4,5). Colleges in the UK and USA have already reported outbreaks among the student population, necessitating closure in some cases (6). The UK government is exploring community-wide testing for asymptomatic COVID-19 infection as a potential health protection tool, to enable outbreaks to be identified and controlled early (7). A SAGE consensus statement has suggested that such a strategy might be useful in “well-defined higher risk settings”, such as universities (8). This method is largely untested within a university setting, however. This pilot study was based in the Norwich Research Park (NRP), which includes the University of East Anglia (UEA) and a range of business and research institutions. The study offered four COVID-19 polymerase chain reaction (PCR) swabs to all staff and students on the site over a two-week period. The aim of the study was to pilot participant guidance materials, logistics, laboratory and data processes and the user-facing web application. It also aimed to establish costs and to assess participant acceptability.

## Methods

All participants living or working on the NRP were eligible to participate and were invited to join the study via an email cascade to staff and students. Ethics approval (no. 2019/20-140) was obtained from the UEA research ethics committee. A secure web application was developed and hosted by the School of Computing Sciences at the UEA. This involved the design and implementation of software to facilitate participant sign-up for booking slots, email authentication and data management protocols to host test results and participant personal data. Participants registered on the web application and were invited to verify their email accounts and sign in using an industry standard authentication protocol. All those who verified their accounts were considered to have enrolled in the study. 180 people participated in a pre-trial, in which they returned two swabs. For the main study, participants were offered four swab tests over two weeks. They collected swab kits, self-administered the swab and returned the test in pre-booked return slots. For the purposes of this report the pre-trial and main trial are considered as a single trial.

Swabs were processed in dedicated laboratory facilities at the Earlham Institute (EI) from Monday to Friday. Copan Liquid Amies Elution Swabs (Eswabs) were used for all participants. Participant samples were tested for the presence of SARS-COV-2 using a quantitative polymerase chain reaction (qPCR) assay. Briefly, nasopharyngeal swab samples were pre-treated with a lysis buffer (Cytiva) that disrupts human cells and viral particles to release nucleic acid into solution. Following inactivation, RNA was extracted using Sera-Xtracta Virus/Pathogen Kit (Cytiva) on a liquid handling platform (Beckman NXp). RNA extracts were amplified for detection of the target genes using a set of optimised primers and probes (2019-nCoV CDC EUA Kit, IDT), and enzymes (qPCRBIO Probe 1-Step Go No-ROX, PCRBioystems) in a real-time PCR system (Quantstudio5, Thermofisher). The assay is qualitative with results assessed based on a threshold cycle (Ct value) to determine outcome (positive, negative, insufficient) using a combination of Ct value for the viral target (N1) and human internal control gene (RPP30) genes. Positive and negative controls were included in every RNA extraction and qPCR run for quality control. Sample data was managed at EI using Exemplar LIMS® from Sapio Sciences. Results data was processed from these samples using Python 3 scripts developed at EI and running on virtual infrastructure provided by the CyVerse UK cloud. Validated participant results were then sent to the information systems at UEA using secure web services.

Negative or inconclusive results were posted on participants’ online accounts, with additional encoding and encryption protocols deployed to maintain data security. A protocol for managing positive results was developed, including notifying participants and NHS Track and Trace by telephone, and advising participants to share their results with their GP. After the completion of the feasibility project, participants who had returned at least one swab were emailed a link to complete a short online participant feedback questionnaire, including questions on demographics and their experience of the project. Groups were compared with Chi-squared tests and free text responses were analysed by extraction of key themes. Participant demographics were summarised in a table, and Chi-squared tests used to compare those who enrolled but did not participate in the study, and those who did participate. Resource use was summarised in a table and the flow of participants and swabs through the study summarised in a flow diagram.

## Results

Results are summarised in Figures 1 and 2 and Tables 1 and 2. 180 people participated in the pre-trial and another 873 in the main trial (Table 1). 458 participated in the post-study survey (Figure 2). 19% of the eligible population enrolled in the study and 24% of these dropped out of the study prior to returning any samples (Figure 1). 86% (687/798) of participants who received at least one result returned all four swabs. 6 participants received 1 inconclusive result. All other results were negative. All participants received at least one negative swab. Participants could choose to return their swabs on foot or by car: pedestrian access sites were favoured over vehicle access sites. There was no lag between the results upload and participants receiving their result notification via email. The post-trial survey found that the overall acceptability rating was 4.5/5 stars (with 5 being the most positive) and 97% of participants would take up repeat testing if available. Self-swabbing received the lowest score for participant acceptability (71% agreed or strongly agreed that taking the swabs was easy to do). 81% of responders to the post-trial survey were staff and 16% were students.

**Table 1.** Participant characteristics.

|  | **Total**<br>**n=1,053<sup>a</sup>** | **Percentage<sup>b</sup>** |
| --- | --- | --- |
| **Sex** |  |  |
| Female | 579 | 55.0% |
| Male | 436 | 41.4% |
| Non-binary | 4 | 0.4% |
| Prefer not to say | 34 | 3.2% |
| **Age band (years)** |  |  |
| 18-24 | 120 | 11.4% |
| 25-34 | 328 | 31.1% |
| 35-44 | 258 | 24.5% |
| 45-54 | 201 | 19.1% |
| 55-64 | 118 | 11.2% |
| 65+ | 28 | 2.7% |
| **Ethnicity** |  |  |
| Asian | 60 | 5.7% |
| Black | 2 | 0.2% |
| Mixed | 31 | 2.9% |
| Other | 22 | 2.1% |
| Prefer not to say | 49 | 4.7% |
| White | 889 | 84.4% |

<sup>a</sup>Total participants who registered for the study and validated their account
<sup>b</sup>There was no significant demographic difference between those enrolled but did not participate and those who did participate.

**Table 2.** Resources.

|  |  |
| --- | --- |
| Laboratory staff time [excluding planning and reporting] | 1,200 hours |
| Total laboratory staff salary costs [excluding planning and reporting] | £19,385 |
| Average sample processing time | 35.5 hours |
| Weekday average sample processing time | 25.5 hours |
| Administrative staff time estimate | 500 hours |
| Laboratory consumables cost estimate per test kit | £3.61 |
| Total number of swabs supplied to participants | 3,360 |
| Incurred cost<sup>a</sup> | £75,906 |
| Estimated in-kind contributions<sup>b</sup> | £89,612 |

<sup>a</sup>This figure represents the spend on services and equipment. The feasibility study inevitably incurred a high unit cost, and for example tripling the test numbers would have a negligible effect on cost.
<sup>b</sup>This figure represents the costs absorbed by the existing service.

**Figure 1.**
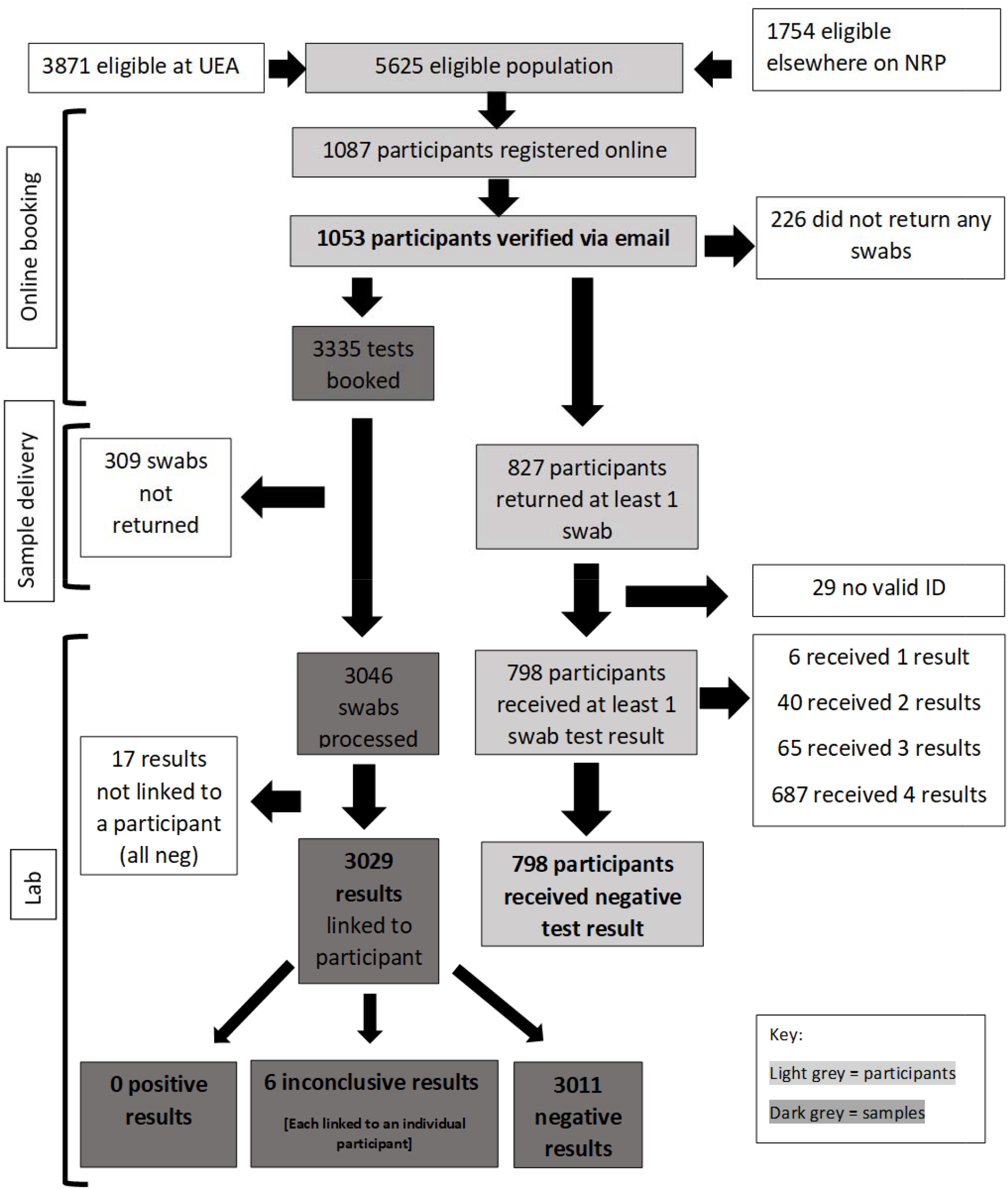
Flow of participants through the study and main results.

**Figure 2.**
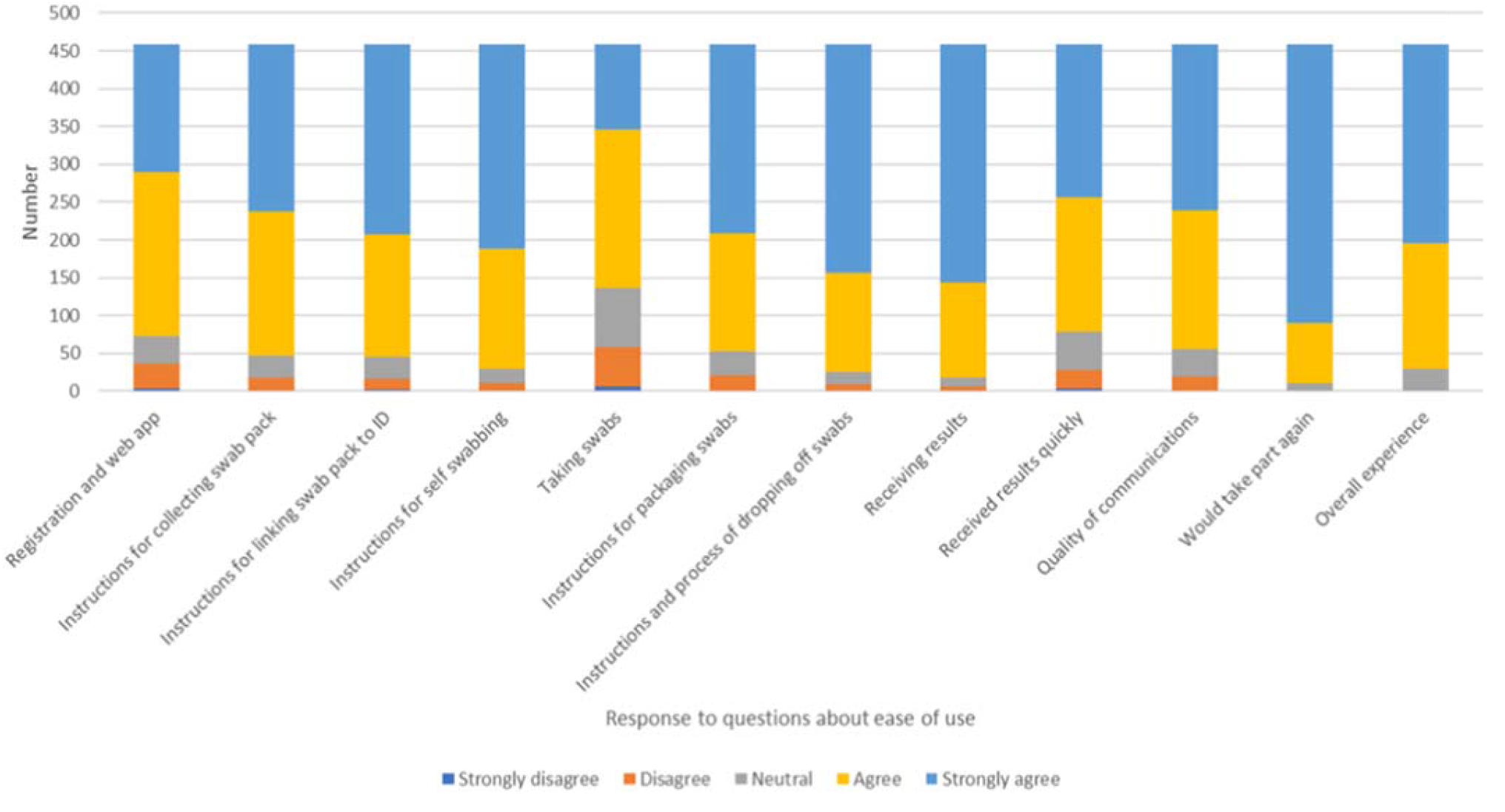
Participant responses.

### Guidance documents

An analysis of guidance documents provided for participants, including an instruction leaflet, standard emails and website text found an average Flesch reading ease score of 60.1. This indicates that material should be comprehensible to a person aged 13-15 years. The instruction leaflet for participants undertaking swabbing had a score of 74, which indicates greater readability. The text for participants opening an online account had a score of 46.8, which indicates that the reader requires a university education to understand the text.

### Participant acceptability

Participant acceptability was assessed in two rounds: first by inviting email comments during the testing process and second by a formal survey. Key themes emerging from participant emails included eligibility and uncertainty about the logistical processes. There were very few emails regarding the process of self-swabbing.

458 participants (57%) responded to the survey, which included 11 questions about the acceptability of the process, with responders choosing their response to each question on a 5-point scale from ‘strongly agree’ (scored as 5) to ‘strongly disagree’ (scored as 1) (Figure 2). Responders were generally positive about their experience of the project, and the overall mean response score was 4.51 stars out of 5 (with 5 being the most positive). 89% of those who responded to the survey returned all four swabs. 34% of responders were working or studying on site, with the rest working either partly or exclusively from home. 77.5% of survey responders lived 5 miles or less from the NRP. Responders were not significantly more likely than all 798 participants to have returned all 4 swabs (89% and 86% respectively, p=0.46). There were no statistically significant differences between those who provided all 4 swabs (n=409) and those who provided fewer than 4 (n=49) in demographics or any answers, including the mean response score (4.54 and 4.50 stars out of 5 respectively, p=0.83). 43 of the 49 responders who provided fewer than 4 swabs gave a reason: 21 (48%) were away during part of the study, and another 8 (19%) forgot or were unexpectedly busy with other commitments.

266 responders (59%) answered the free text questions (“Is there any feedback you would like to share about any aspect of your participation in the project?” and “Is there a reason why you were unable to take any self-swab samples?”). Responses were generally positive and included requests for ongoing testing, feedback on results of the study and praise for organisation and response to enquiries. Responders recommended clearer communication on the variability of time to receive results, as some interpreted a longer wait as being suggestive of a positive result. They also provided useful feedback on the usability of the web application, particularly the sign-up process: they requested a simpler approach, and recommended changes to the presentation of results to reduce anxiety. Responders also requested clearer instructions regarding packing samples.

## Discussion

### Main findings of this study

827 participants took and delivered their swabs over a two-week period. The relatively low uptake can be explained by the timing of the study during the summer break, the absence of staff from campus due to working-from-home policies and a short run-up to the study. Nearly a quarter of participants dropped out of the study prior to returning any samples. The reasons for this were unclear from the evaluation, however there was no significant demographic difference between those who enrolled in the study but did not participate, and those who did participate.

The sex distribution of the eligible population was not available, it is therefore not possible to determine whether the sample population (55% women) was reflective of the eligible population. The ethnic distribution of the study population was broadly reflective of the population of Norwich: 10.9% of participants were of Asian, black, mixed or ‘other’ ethnicity, compared to 9.1% for Norwich (9).

The study did not identify any false positives, despite this being considered a risk of universal testing (10). An analysis of guidance documents and participant emails indicates a need for clearer information tailored to the eligible population. The participant questionnaire revealed a high level of participant engagement and acceptability. Combined with the low drop-out rate after taking the first swab (86% of participants who returned at least one swab returned all four), this suggests that participants found self-swabbing and the collection and delivery of samples generally acceptable.

The laboratory and web application processed 3046 swabs during the study. Laboratory processes were efficient, with an average processing time of just over 24 hours during the week. The reagent cost per test of £3.61 was based on a batch size of 6,000 tests which were ordered quickly from outside the UK, and we would expect lower costs per test in a larger initiative. The secure web application was hosted by the School of Computing Sciences at minimum cost. Similarly, the computing infrastructure at EI was provided in kind by CyVerse UK at minimal cost to the project.

### What is already known on this topic

The evidence base for use of asymptomatic testing for COVID-19 as an infection control measure remains limited. Universal, repeat testing has been advocated however, as a means of avoiding lockdown (11). Universities across the world are now considering universal testing despite the pitfalls of this strategy, which include false positive and negative tests, the difficulty of defining an active infection and significant cost (10,12). UK universities and colleges in the USA have already reported outbreaks of COVID-19 (12). The potential for COVID-19 transmission in universities is significant, particularly shortly after the beginning of term when students return to campus (4,5). Models demonstrate that universal testing may have a significant impact on control of the virus, depending on the ability of the setting to implement other control methods (13). There is however, no published study assessing the feasibility or acceptability of a universal programme for COVID-19 testing on a university campus.

### What this study adds

This pilot study indicates that universal testing on a university campus is both feasible and acceptable to the population. A strength of this study is that it trialled the feasibility of repeat testing for COVID-19 in a relatively large, asymptomatic population within a research park and university campus. Participants included both staff and students and the findings can be applied both to a larger study on the same site and to other university contexts. There was a high level of participant engagement with the study. This study has demonstrated that clear, consistent communications and an intuitive web application are necessary for helping participants to understand the need for testing and the process of undertaking and returning the test.

Both universal testing and the current UK national public health strategy of testing symptomatic people via a local testing site have strengths and weaknesses. The current national strategy of symptomatic testing is adequate when there are few cases in the community, and is cheaper in the short term, but risks allowing undetected spread of COVID-19 when cases start to rise in a community, particularly when results take more than 24 hours to be reported. The main potential problem with universal testing is that it may generate false positives, and therefore unnecessary contact tracing and isolation. It is also more expensive in the short term. There were no false positives out of 3,046 tests in this study. The main advantage of universal testing is that it can identify infectious asymptomatic cases and isolate them before they can infect others in the community. This is a major benefit on a campus university with large numbers of students in a community where isolation and social spacing may be challenging to maintain, and where a major outbreak would have serious consequences for students’ education, the university, and the local community.

### Limitations of this study

Limitations of the study include the relatively low uptake and the low prevalence of COVID-19 in this population, which meant that processes for managing positive results could not be tested. At the time of the study, community prevalence of COVID-19 was approximately 1 in 1700 people(15). As this was a self-selecting cohort of university staff and students, motivation to participate may be higher than in the general population. The findings are generalisable to university staff but may be less generalisable to new undergraduates. This study used PCR swabs but acceptability of some alternative testing methods, such as saliva testing, may be even higher.

## Data Availability

The datasets generated during during the current study are available from the corresponding author on reasonable request.

## Acknowledgments

Many thanks to the study participants. Thanks also to the volunteers who assisted with this project and members of the Norwich Covid-19 Testing Initiative (Jose Carrasco-Lopez, Leah Catchpole, Kirsty Culley, Fiona Fraser, Geoff Plumb, Chris Watkins).

## Contributors

TBG, NSt, and JC wrote the first drafts of this manuscript. EC, NH, RD, DE, MH, KG, SC, GJ, EA PB, NSt and TBG were involved in the conception and design of the project. KG, NSh, TB, RD, SR, TOB, SH, NI, GP, AD and JL developed and implemented laboratory processes. SC, JSC, GP, RS, EA, GJ and PB contributed to data collection. MH, SC, NH and DE oversaw and managed the project. TBG, NSt, and JC reviewed and analysed the results. All authors contributed to or reviewed the manuscript.

